# Comparison of infliximab versus cyclophosphamide on treatment of refractory uveitis of Behcet’s disease: pilot clinical study

**DOI:** 10.1101/2024.12.11.24317445

**Authors:** Alireza Sadeghi, Arezoo Karimimoghadam, Fereydoun Davatchi, Farhad Shahram, Saeideh Mazloomzadeh, Azadeh Behnamghader, Nooshin Jalili, Sajjad Biglari, Zahra Jourahmad

## Abstract

**Background:** To evaluate the effect of infliximab (IFX), in comparison with cyclophosphamide (CP) on refractory uveitis of BD.

**Methods:** Subjects with uveitis and retinal vasculitis (RV) resistant to conventional treatment were included. Ten patients were divided in the IFX and CP groups. Patients in the IFX group received four courses of 3 mg/kg of IFX, intravenously. Patients in the CP group received four courses of pulse CP of 1000 mg, by the monthly interval. All patients received weekly 25 mg of methotrexate, (or 5 mg/kg of cyclosporine daily), and 3 mg/kg and 1 mg/kg of daily azathioprine and prednisolone, respectively. To evaluate the treatment effects, visual acuity (VA), anterior uveitis (AU), posterior uveitis (PU), and RV were assessed, at the baseline and four months after treatment.

**Results:** In ten patients included in this study there were no significant differences in the treatment effects of IFX, compared to the CP regarding VA, PU, AU and RV. RV became inactive after four months of treatment in all five patients in the IFX group versus four patients in the CP group.

**Conclusion:** IFX and CP have the same efficacy in the short-term treatment of uveitis in BD. In future investigations, multi-center studies with larger sample sizes are required to investigate the cost-effectiveness of IFX in the treatment of uveitis of BD.

**Highlights:**

- A new anti-TNF biologic agent, infliximab and cyclophosphamide have the same efficacy in the treatment of uveitis in BD, and since it has less toxicity, it is a superior treatment option.
- In areas where access to the contemporary treatment is limited and expensive, the use of traditional medication results in an increased quality-adjusted life year (QALY).
- Multi-center studies with larger sample sizes are required to investigate the cost-effectiveness of infliximab in the treatment of uveitis of BD.

## 1. Introduction

Behcet’s disease (BD) is a multisystem disease with an underlying inflammatory mechanism.^1^ The corticosteroids (CSs) are the main treatment of Behcet’s uveitis. The combination of CSs, with conventional immunosuppressive (CIS), including azathioprine, cyclosporine-A, and methotrexate are the second-line treatments for ocular involvement in BD.^2^ However, there is no firm evidence to suggest the best treatment regimen in uveitis refractory to these agents. The immunomodulatory therapy (IMT) for uveitis includes antimetabolites, inhibitors of T-lymphocytes signaling, biologic response modifiers (BRMs), and alkylating agents. If the uveitis is severe and refractory the more potent immunomodulatory therapy is needed. Cyclophosphamide (CP) an alkylating agents has for many years showed efficacy and are traditionally considered in these cases.^3^ CP has showed the most effective immunosuppressive treatment for uveitis in patients at risk of vision loss due to inflammatory diseases.^4^ BRMs including interferon-_α_, anti-TNF-_α_ antibodies, and IL-1 blocking agents are newer drugs introduced for the treatment of inflammatory uveitis. In this group, infliximab (IFX) —an anti-TNF-_α_ antibodies— is specially used in many randomized clinical trials alone or in combination of other drugs and showed promising results in maintaining visual acuity in refractory cases.^5^ In this pilot study, we aimed to evaluate the treatment effects of IFX, in comparison to CP in refractory uveitis of BD. Our study is the first to compare the treatment effects of IFX and CP on intractable uveitis in BD.

## 2. Methods

### 2.1. Patients’ selection

In this single-blind pilot study, we recruited ten patients with BD and treatment-resistant ophthalmic lesions, who were referred to the rheumatology clinic of the University of Medical Sciences. The patients were selected according to the International Criteria for Behcet’s Disease (ICBD).^6^

The patients were divided into two groups of five patients. The first group, received IFX plus prednisolone, azathioprine, and methotrexate (or cyclosporine) and the second group received CP plus prednisolone, azathioprine, and methotrexate (or cyclosporine), as a treatment regimen. Despite the growing number of publications for treatment of refractory uveitis, there is no consensus for the definition of refractory uveitis in general. The majority of studies consider clinical variables for the definition of refractory uveitis.^7^ In this study the inclusion criteria were subjects with the diagnosis of well-established uveitis, retinal vasculitis (RV), or macular edema based on ophthalmology examination and fluorescein angiography who were resistant to conventional treatment (1 mg/kg of prednisolone daily with 3 mg/kg of azathioprine daily and 5 mg/kg cyclosporine daily/or 25 mg of methotrexate weekly) and more than two score drop in VA for at least three months.

This study was approved by The Regional Bioethics Committee of the University of Medical Sciences. The patients entered the study voluntarily. The written consent form was acquired from all the patients, and they were informed of the possible medication side effects. Patients were monthly followed up for the appearance of any treatment-related adverse events, and in the case of any emergence of complication, the patient was excluded from the study.

### 2.2. Treatment

In the IFX group, patients received 3 mg/kg of IFX, intravenously at the beginning and then, at the 2nd, 6th, and 8th week. Also, they received 25 mg of methotrexate weekly (or 5 mg/kg cyclosporine daily), and daily 3 mg/kg, and 1 mg/kg of azathioprine and prednisolone, respectively. In the CP group, patients received pulse CP of 1000 mg/per month, for 4 months, 25 mg of methotrexate weekly (or 5 mg/kg cyclosporine daily), and daily 3 mg/kg and 1 mg/kg of azathioprine and prednisolone, respectively.

### 2.3. Ophthalmologic evaluation

All the patients received ophthalmologic examinations during the monthly follow-up throughout the study. The examination findings were gathered through a predefined questionnaire by a retina specialist, who was blinded to the treatment. To evaluate visual acuity (VA) we used the Snellen chart. We evaluated anterior uveitis (AU), posterior uveitis (PU), and RV, as the main parameters of uveitis, due to BD. These parameters were graded and scored separately, using the BenEzra scoring system ^8^ at the beginning of the study, then at the first, second, third, and fourth months of treatment. The benezra uveitis scoring system is designed to determine the disease activity in uveitis and to evaluate the treatment effects of anti-inflammatory and immunomodulating drugs. This system assesses the changes in five components of eye separately (anterior segment, vitreous, fundus, visual acuity, and fluorescein angiography) and according to quantitative evidence assigns a score from 0 to 5. The fluorescein angiography was used for all patients at the beginning and at the end of the study. The presence of RV was assessed monthly by a retina specialist using slit-lamp biomicroscopy with 90-diopter lens.

### 2.4. Statistical analysis

For statistical analysis we used IBM SPSS version 16 software (SPSS Inc, Chicago, IL, USA). The normality of data was examined using Shapiro-Wilk test. To evaluate the treatment effect of IFX and CP we used Friedman test and to compare the between group efficacy of two drugs Mann Whitney U test was applied. The between group changes in RV was evaluated using Fischer Exact test. The p-value less than 0.05 was deemed to be statistically significant.

## Results

A total number of 10 patients (6 males and 4 females) participated in this study. The mean patients’ age was 34.8±5.3 years (37.0±11.8 years in IFX group and 28.6±10.31 years in CP group). The follow-up time was four months. The patients’ demographics are demonstrated in Table 1.

**Table 1.**
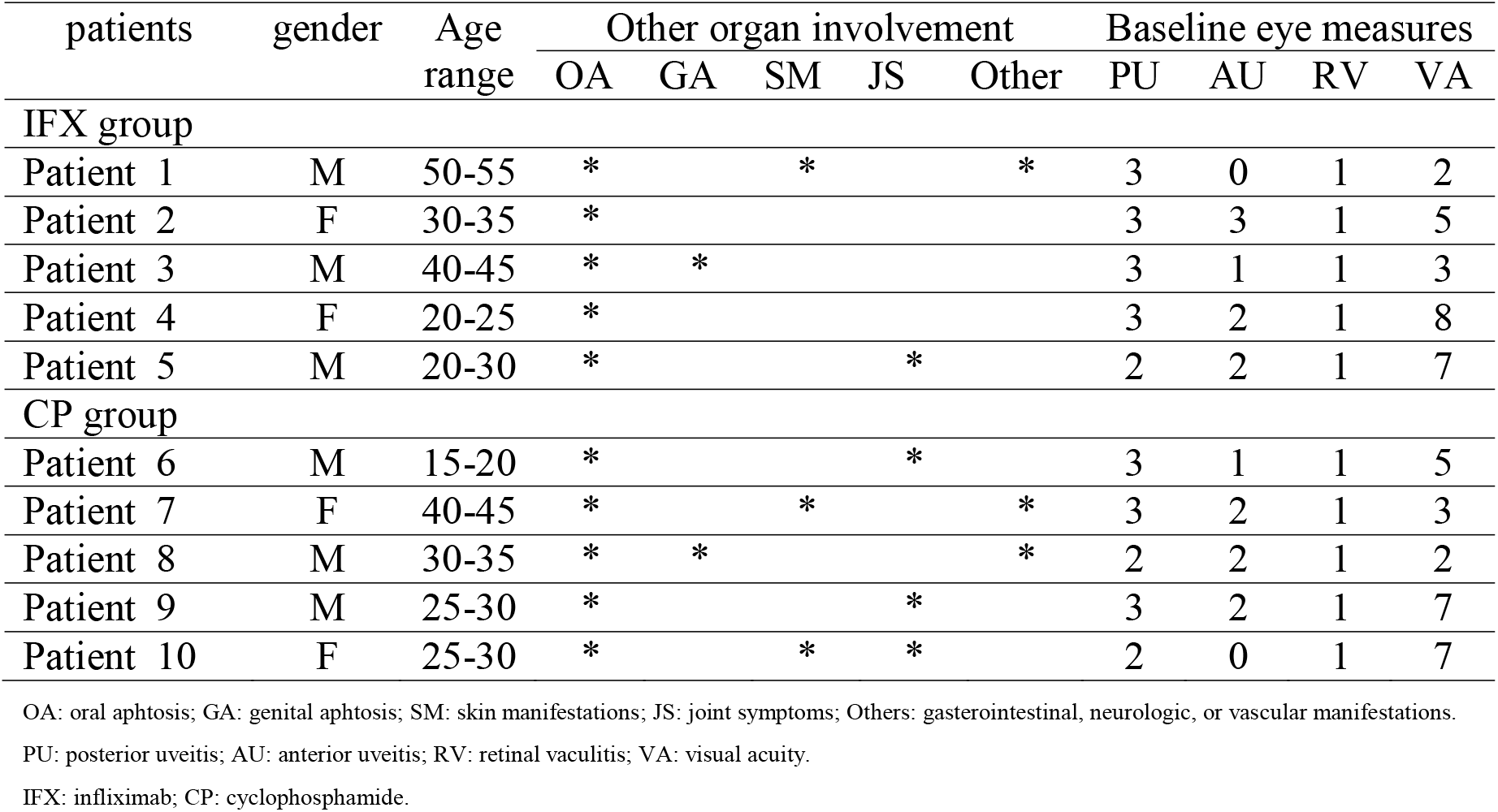
Patients’ characteristics.

Regarding the baseline measures the RV was present in all patients, the mean AU was 1.5±0.972, the mean PU was 2.7±0.483, and the mean VA was 4.9±2.28. There were no statistically significant differences in the mean of PU, AU or VA between the two groups. There was a significant improvement in the mean VA, AU, and PU from baseline in both treatment groups at the fourth month of the study (Table 2). The mean VA improved from the mean of 5.0 to 8.6 in the IFX group (p-value=0.002) and from the mean of 4.8 to 9.0 in the CP group (p-value=0.001). Also, the mean PU improved significantly from the mean of 2.8 to 0.00 in the IFX group (p-value=0.001), and from 2.6 to 0.4 in the CP group (p-value=0.001). As well, the mean AU improved significantly from 1.6 to 0.00 in the IFX group (p-value=0.009) and from the mean of 1.4 to 0.00 in the CP group (p-value=0.012) at the fourth month of study. The details are shown in Table 2. There were no statistically significant differences in the mean of VA, AU, and PU between the groups (Table 3).

**Table 2.** Data of patients’ eye during follow-ups.

| Groups |  | Mean±SD |  |  |  |  | p-value |
| --- | --- | --- | --- | --- | --- | --- | --- |
|  |  | baseline | 1<sup>st</sup> month | 2<sup>nd</sup> month | 3<sup>rd</sup> month | 4<sup>th</sup> month |  |
| IFX | AU | 1.60±1.14 | 1.00±0.70 | 0.40±0.54 | 0.20±0.44 | 0.00±0.00 | 0.009 |
|  | PU | 2.80±0.44 | 2.20±0.44 | 1.40±0.54 | 0.60±0.54 | 0.00±0.00 | 0.001 |
|  | VA | 5.00±2.55 | 5.00±2.55 | 6.00±2.00 | 7.60±2.19 | 8.60±2.07 | 0.002 |
| CP | AU | 1.40±0.89 | 1.20±1.09 | 0.80±0.83 | 0.20±0.44 | 0.00±0.00 | 0.012 |
|  | PU | 2.60±0.54 | 2.40±0.54 | 1.40±0.54 | 0.60±0.54 | 0.40±0.54 | 0.001 |
|  | VA | 4.80±2.28 | 6.40±1.81 | 7.80±2.16 | 8.20±2.38 | 9.00±2.23 | 0.001 |

\*measured by Friedman test
PU: posterior uveitis; AU: anterior uveitis; VA: visual acuity; IFX: infliximab; CP: cyclophosphamide, SD: standard deviation.

**Table 3.** Data of patients’ eye compared in two groups.

|  |  | VA |  | PU |  | AU |  |
| --- | --- | --- | --- | --- | --- | --- | --- |
| Groups |  | Mean±SD | P-value | Mean±SD | P-value | Mean±SD | P-value |
| IFX | Baseline | 2.55±1.14 | 0.841 | 2.80±0.44 | 0.690 | 1.60±1.14 | 0.841 |
| CP |  | 2.28±1.02 |  | 2.60±0.54 |  | 1.40±0.89 |  |
| IFX | 1<sup>st</sup> month | 5.00±2.55 | 0.421 | 2.20±0.44 | 0.690 | 1.00±0.70 | 0.690 |
| CP |  | 6.40±1.81 |  | 2.40±0.54 |  | 1.20±1.09 |  |
| IFX | 2<sup>nd</sup> month | 6.00±2.00 | 0.095 | 1.40±0.54 | 1.000 | 0.40±0.54 | 0.548 |
| CP |  | 7.80±2.16 |  | 1.40±0.54 |  | 0.80±0.83 |  |
| IFX | 3<sup>rd</sup> month | 7.60±2.19 | 0.548 | 0.60±0.54 | 1.000 | 0.20±0.44 | 0.548 |
| CP |  | 8.20±2.38 |  | 0.60±0.54 |  | 0.20±0.44 |  |
| IFX | 4<sup>th</sup> month | 8.60±2.07 | 0.421 | 0.00±0.00 | 0.310 | 0.00±0.00 | 1.000 |
| CP |  | 9.00±2.23 |  | 0.40±0.54 |  | 0.00±0.00 |  |

\*measured by Mann Whiteny U test
PU: posterior uveitis; AU: anterior uveitis; VA: visual acuity; IFX: infliximab; CP: cyclophosphamide, SD: standard deviation.

There was an improvement in the RV score in 100% of patients in the IFX group, and 80% of the patients in the CP group; however, this difference was not statistically significant. Table 4 shows the details of changes in the RV score from the baseline in two groups, throughout the study. There was not any report of serious adverse events within the four months follow-up of patients during the study.

**Table 4.** RV changes from base line in first, second, third, and forth months of treatment in two treatment groups.

| changes from baseline | RV | Frequency |  |
| --- | --- | --- | --- |
|  |  | IFX group (N=5) | CP group (N=5) |
| Baseline | Present | 5 | 5 |
|  | Resolved | 0 | 0 |
| First month | Present | 5 | 5 |
|  | Resolved | 0 | 0 |
| Second month | Present | 4 | 5 |
|  | Resolved | 1 | 0 |
| Third month | Present | 3 | 4 |
|  | Resolved | 2 | 1 |
| Forth month | Present | 0 | 1 |
|  | Resolved | 5 | 4 |

\*measured by Fisher Exact Test for cell with 0 frequency
RV: retinal vasculitis; IFX: infliximab; CP: cyclophosphamide; SD: standard deviation.

## Discussion

In this study, we evaluated and compared the treatment effects of IFX and CP on the refractory uveitis in patients with BD. VA, AU, PU, and RV were evaluated before the study and during the first, second, third, and fourth months after treatment. Our results showed that there was no significant difference in the treatment efficacy between the two groups; however, within-group analysis, comparing the parameters before the study and four months after the treatment, showed that both medications could ameliorate VA, AU, and PU scores significantly from the baseline. In addition, our results showed that both drugs prevented the progress of RV at the end of the study.

BD is a complex multisystem disease with a proposed inflammatory underlying cause. Therefore, strategies that target the immune system involved in BD’s pathophysiology, are effective in the treatment of disease or ameliorating the severity of complications. Therefore, the class of drugs that target the immune system could be an appropriate treatment choice.^**9**^ Immunosuppressive drugs including antimetabolites, T-cell inhibitors, and alkylating agents are drugs with immunomodulatory mechanism, which are used in the treatment of ocular inflammatory diseases.^**10**^ CP is an alkylating agent that by depleting immunocompetent lymphocytes works as an immune suppressor in the treatment of autoimmune diseases, including uveitis in BD.^**11**^ In previous studies, CP is shown to be effective in the treatment of uveitis in BD, particularly in severe cases of uveitis, such as posterior uveitis and retinal vasculitis, showing resistance to other treatments.^**4,12–14**^

Compared to other immunosuppressive drugs, CP induces disease remission in most of the patients; however, major side effects such as myelosuppression has caused limitation to be used only in the severe case of ocular inflammation.^**4**^ Due to the accelerated drug action, and fewer side effects, as well as the more specific mechanism of action against inflammatory processes, biologic agents have become a superior alternate to immunosuppressive drugs in the treatment of autoimmune uveitis.^**15**^

Uveitis, as a sight-threatening disease is the main reason for morbidity in BD. The breakthrough of biologic agents such as anti-TNF-_α_ antibodies has improved the visual prognosis in patients with BD significantly.^16^ Anti-tumor necrosis factor (anti-TNF)-biologic agent, including infliximab (IFX), is suggested by the European League against Rheumatism (EULAR) for the treatment of intractable uveitis of BD.^17^ The alkylating agents, such as cyclophosphamide (CP) induce remission and are effective treatment for intractable uveitis of BD; however, because of their toxicity are mostly used for life-threating complication such as major vessel involvement.^18^

IFX is a monoclonal antibody against TNF-_α_ and exerts its anti-inflammatory effects, through induction of programmed cell death in T-lymphocytes and macrophages.^**19**^ Uveitis due to BD can affect anterior, intermediate, and posterior segments.^**20**^ IFX alone or in combination with conventional drugs has shown to be effective in the treatment of all types of uveitis and RV resistant to treatment in BD.^**21,22**^ Moreover, the comparison of IFX versus conventional treatments of uveitis in BD, in the previous studies has shown the superiority of IFX.^**23–26**^ Previous studies have separately addressed the efficacy of IFX and CP on ocular involvement, in the course of BD. Our study is the first of its kind to investigate the efficacy of IFX, in comparison to CP on intractable uveitis in BD. Our results are in line with previous studies and showed that IFX, a biologic drug is an effective treatment for intractable uveitis of BD. Side effects of IFX, compared to CP are milder and include autoimmune diseases, increased risk of infection, and injection site reaction,^**27**^ versus bone marrow suppression, increased risk of malignancies, alopecia, nausea, and vomiting.^**28**^ In this pilot study, after four months of treatment, none of the patients reported any side effects with the two drugs. In addition, we showed that IFX is as efficacious as CP for the treatment of intractable uveitis in BD and due to the fewer side effects could be an appropriate replacement for CP. Moreover, some studies showed the benefits and encouraged the earlier use of IFX, compared to CISs for uveitis in BD.^**5,29**^ There are still ongoing studies to evaluate the treatment effects and superiority of IFX, compared to conventional drugs in the treatment of uveitis in BD.

Visual prognosis in patients with Behcet’s uveitis could be assessed based on visual acuity. In an international study to compare the ocular features of BD in 14 countries, poor vision was reported significantly more frequent in India, Iran, and Japan.^**30**^ Therefore, we recommend physicians in these regions to start combination therapy with more potent immunomodulatory agents earlier in order to have a better visual outcome and avoiding vision loss.

Currently, the market for monoclonal antibodies is primarily in developed countries. These treatments are not registered for use in low-income countries and in middle-income countries they are not provided by the public health systems and are only available restrictively at a high cost through private sectors.^**31**^

In Iran, the cost of infliximab is ten times that of cyclophosphamide and is not covered by public insurance, making it less accessible to a large proportion of the patient population in the country.

The most important limitations of our study are due to the study design, short study time, and small sample size; however, our results can be cited as a preliminary study to guide more broad projects that weight the cost-effectiveness of IFX to CP, in the treatment of intractable uveitis in BD.

## Conclusion

In conclusion, this study showed that IFX and CP can both effectively treat intractable uveitis in patients with BD, there is no significant difference in efficacy between the two drugs in short-term. Our study highlights the significance of traditional medication as compared to contemporary treatment regimen, particularly in low-income areas where access to medication is limited and costly.

## Data Availability

Data and analysis relevant to the findings of this study can be made available upon request.

## Acknowledgement

This article is resulted from a thesis registered at the University of Medical Sciences.

## Ethical issues

This study was approved by the Ethics Committee for Research on March 28, 2016, with the Code of Ethics (ZUMS.REC.1395.233). The protocol was registered on National Registry of Clinical Trials Website (IRCT.IR) under the ID: RCT20180110038297N1

## Authors contributions

Conceptualization: A S, A K, F D, F S; Methodology: S M; Investigation : A S, A K, F D, F S, N J; Formal analysis and investigation: Z J, A B, S B; Writing - original draft preparation: Z J, S B; Writing - review and editing: F D, F S, A K, N J, S M; Funding acquisition: A B; Resources: A B, N J; Supervision: A S; Approving final version : all authors.

